# Sensitivity and specificity of prediction models based on gustatory disorders in diagnosing COVID-19 patients: a case-control study

**DOI:** 10.1101/2020.05.31.20118380

**Authors:** Kamil Adamczyk, Michał Herman, Janusz Frączek, Robert Piec, Barbara Szykuła-Piec, Artur Zaczyński, Rafał Wójtowicz, Krzysztof Bojanowski, Ewa Rusyan, Zbigniew Król, Waldemar Wierzba, Edward Franek

**Author notes:** Correspondence to: Edward Franek, MD Department of Internal Medicine, Endocrinology and Diabetology (transformed to a COVID-19 department) Central Clinical Hospital of the Ministry of the Interior and Administration in Warsaw ul. Woloska 137 02-507 Warsaw, Poland e-mail address, 0000-0002-5953-7908. Michał Herman. Janusz Frączek. Barbara Szykuła-Piec. Artur Zaczyński. Rafał Wójtowicz. Zbigniew Król.

## Abstract

**Objective:** to quantitatively assess disturbances of sweet, sour and salty and bitter tastes in a group of young, asymptomatic or oligosymptomatic COVID-19 patients; establish a reliable, sensitive and specific test that can diagnose COVID-19 on the basis of taste disorders and publish the results according to STARD 2015 statement (Standards for Reporting Diagnostic Accuracy).

**Design:** case-control study

**Setting:** isolated rooms in the Central Clinical Hospital of the Ministry of the Interior and Administration in Warsaw (which had been transformed into an infectious hospital) and a dormitory in one of Warsaw universities.

**Participants:** 52 SARS-CoV-2 positive (51 men, mean age 21.7 years) and 36 negative students (34 men, mean age 20.8 years).

**Main outcome measures:** a gustatory function assessment (sweet, salty, sour and bitter taste), with flavour concentrations established previously in healthy subjects was conducted for all subjects. Each participant received one tasteless reference and nine flavour tablets with sucrose concentrations of 40, 80 and 106.4 mg/ml; NaCl at 13.5, 17 and 27 mg/ml; ascorbic acid at 6.25 and 12.5 mg/ml and grapefruit extract at 40 mg/ml.

**Results:** the only taste that was impaired significantly more frequently in COVID-19 patients was the sweet taste at the lowest flavour concentration (40 mg/ml, p = 0.002). Different screening and diagnostic models were constructed using the examined variables. The highest accuracy screening test consisted of the positive result of a three-question questionnaire (self-reported loss of taste, self-reported loss of smell, or fever within the last month (positive if at least one present) and/or ageusia of sweet taste at a sucrose concentration of 40 mg/ml. The sensitivity of the model was 94% with a specificity of 55%. The highest accuracy diagnostic test consisted of ageusia of sweet taste at a sucrose concentration of 106.4 mg/ml or/and ageusia of salty taste at an NaCl concentration of 13.5 or 17 mg/ml. The specificity of the test was found to be 100%, and the sensitivity was 34%.

**Conclusion:** as the most effective way of controlling the present pandemic involves testing the wider population for symptomatic, oligosymptomatic and asymptomatic carriers of SARSCoV-2 and isolating or hospitalising infected subjects, in the present study, an inexpensive, simple, fast and sensitive (94%) screening test that can be used for such a purpose is proposed. In addition, a specific (100%) diagnostic test that could be used to refer patients to quarantine in the case of limited availability of genetic or serological tests is proposed.

## Introduction

The novel coronavirus SARS-CoV-2 (severe acute respiratory syndrome coronavirus 2) is the cause of an ongoing pandemic. The outbreak of the novel coronavirus was first reported in the city of Wuhan in Hubei Province, China but rapidly went on to affect almost all countries worldwide.^1^ The disease is characterised by fever, dry cough, dyspnea, chest pain and, in many cases, pneumonia; however, many other non-respiratory signs and symptoms have been described, such as general weakness, myalgia and arthralgia, or headache.^2^

COVID-19 may also have neurological manifestations, some of which may be due to the fact that coronaviruses enter the central nervous system via the peripheral nerves or olfactory bulb and olfactory sensory neurons.^3^ The latter route of viral invasion may be associated with smell disorders, which appear to be highly common in COVID-19 patients. For example, Moein et al. described smell impairment assessed with a well-validated test in 98% of patients, among whom 58% were either anosmic or severely microsmic,^4^ the frequency of self-reported olfactory disorders may be however lower. In a relatively large cohort of a multicentre European study, the frequency of self-reported olfactory dysfunction was 88%.^5^ In another study, the self-reported frequency of anosmia was 67.8% compared with 16% of SARS-CoV-2- negative subjects.^6^

Other non-respiratory manifestations of COVID-19 include signs and symptoms arising from the gastrointestinal tract, such as nausea, vomiting, diarrhoea and anorexia. Such symptoms have been found in a significant percentage (from below 10% to almost 70%) of patients.^7^ Taste disorders have also been reported, and although some reports regard single patients only, dysgeusia and ageusia appear to constitute two common symptoms of COVID-19, occurring in 10% to almost 90% of patients.^5,8,9,10^ However, similar to olfactory dysfunction, most studies regarding taste disorders in SARS-CoV-2 positive patients published to date have been based on self-reported symptoms, and only one study has quantified gustatory dysfunction in patients, though not in healthy control subjects.^11^

Therefore, the present study aimed to conduct a more precise assessment of gustatory function in COVID-19 patients. Specifically, disorders in sweet, sour, and salty and bitter tastes in a group of young, otherwise asymptomatic or mildly symptomatic subjects were quantitatively assessed and compared with a properly matched SAR-CoV-2-negative control group.

In addition, the sensitivity of the relatively expensive RT-PCR test in COVID-19 diagnosis, which is the gold standard diagnostic method for SARS-CoV-2 according to the World Health Organization (WHO) may be may be from different reasons (e.g. shallow swab instead of deep nasopharyngeal one) insufficient,, particularly for single tests.^12,13^ The sensitivity of other diagnostic algorithms using CT scan images may be much higher; however, this procedure is also expensive, and x-ray exposure may lead to adverse effects.^14^ Both the above-mentioned approaches are time-consuming and are not point-of-care procedures. As a result, their suitability for large-scale disease screening is limited. Therefore, this study aimed to establish a reliable, sensitive and specific test to diagnose COVID-19 by assessing taste disorders. The publication was prepared according to the Standards for Reporting of Diagnostic Accuracy Studies (STARD 2015).^15^

## Material and Methods

### Pilot study to determine flavours concentrations

The study aimed to select flavours concentrations to be used in the main study with COVID-19 patients, in order to shorten the researchers’ exposure time and to ensure that previously validated flavour concentrations were applied. It was decided that the weakest taste concentration perceptible to a minimum of 90% of the healthy people surveyed would be chosen, and lower concentrations would be rejected. The highest concentrations of flavours were those shown to be supramaximal stimuli (i.e., subjects could not distinguish the intensity of the taste above the concentrations).

This study was conducted on 25 young, healthy male adults with a median age of 21.11 years (min 19, max 26) with no concomitant diseases, who tested negative for SARS-CoV-2 according to the PCR results of a nasopharyngeal swab test. Each participant received 20 tastant tablets (five different concentrations of four flavours: sour, sweet, salty and bitter). The participants were instructed to rinse their mouths with water. After placing a tastant tablet in the mouth, the participants were instructed to wait until it had dissolved and then describe the perceived taste. The participants did not know which flavour tablets they had received. Each participant was asked to estimate the flavour and strength of each tastant tablet on a scale of 1 to 5.

Water was used as the carrier substance (filler) and, after dissolving a flavour, gelatine and agar were added to enable tablets to be formed. Each tablet had an identical volume. The tablets contained different flavours in the following concentrations:

- Five concentrations of a sour taste (ascorbic acid): 3.125, 6.25, 12.5, 18.75, 25 mg/ml
- Five concentrations of a sweet taste (sucrose): 20, 40, 60, 80, 106.4 mg/ml
- Five concentrations of a salty taste (sodium chloride): 7.75, 13.5, 17, 27, 34.75 mg/ml
- Five concentrations of a bitter taste (grapefruit extract): 20, 30, 40, 50, 60 mg/ml

During this pilot study, difficulties in identifying the bitter taste were observed, and it was often confused with the salty taste. Therefore, we decided to reject the diagnostic values of various bitter taste concentrations, which left only the lowest perceptible concentration.

Finally, based on the above criteria, the concentrations with the highest diagnostic values were selected from the set of the previously discussed concentrations, along with the names assigned to the tastant tablets:

1. Sour:

a. Sour 1 – 6.25 mg/ml
b. Sour 2 – 12.5 mg/ml
2. Sweet:

a. Sweet 1 – 40 mg/ml
b. Sweet 2 – 80 mg/ml
c. Sweet 3 – 106.4 mg/ml
3. Salty:

a. Salty 1 – 13.5 mg/ml
b. Salty 2 – 17 mg/ml
c. Salty 3 – 27 mg/ml
4. Bitter:

a. Bitter 1 – 40 mg/ml

### Main study

The main study was conducted from April to May 2020 on 92 students from Warsaw, Poland (all of those living in a dormitory) who fulfilled the inclusion (confirmed SARS-CoV-2 infection, aged over 18) and exclusion criteria (lack of informed consent, smell or taste disturbances lasting more than three months on the day of the test, any neurological disease, any disease or factor that in the opinion of an investigator may influence the tests) and who provided signed informed consent. Due to the epidemiological situation, after detecting a SARS-CoV-2 coronavirus outbreak in one of the Universities in Warsaw, Poland, students living together in the dormitory were tested for SARS-CoV-2 using the RT-PCR test of nasopharyngeal swab, according to the WHO standards. Students with a positive test result were isolated from those with negative test results and were transferred to isolation rooms in a specially adapted hotel. Students who tested negative were placed in individual rooms in the school dormitory. Students from both groups were placed under the supervision of the Central Clinical Hospital of the Ministry of the Interior and Administration in Warsaw, which had been transformed into an infectious hospital.

Eighty-eight students completed the study. Of these students, 51 were in the study group (100% included in the study) and 37 were in the control group (90% included in the study; four study subjects did not return the questionnaire). The study flow is shown in Figure 1. On the day of the examination, an additional swab was collected, and a PCR test performed in each student to confirm SARS-CoV-2 infection. Therefore, at the time of the data collection, the researchers knew the results of the first PCR test but not the second. Only one person from the control group tested positive for SARS-CoV-2 after re-examination. This result was confirmed with a PCR test, and the participant was transferred from the control group to the test group (therefore, the final number of participants in the control group was 36 and the study group was 52) for further statistical analysis. RT-PCR tests were conducted in the National Institute of Public Health - National Institute of Hygiene (NIPH-NIH), which is the main governmental public health research institute and the reference centre for the national network of sanitary epidemiological service.^16^ The GeneFinder COVID-19 PLUS RealAmp Kit was used, detecting RdRp, E and N genes, with a sensitivity of 10 copies of these genes, and cut-off point of ≤43 cycles for a positive result. The assessors of the PCR (reference) tests were not aware of any clinical information regarding participants or the flavour test results.

**Figure 1.**
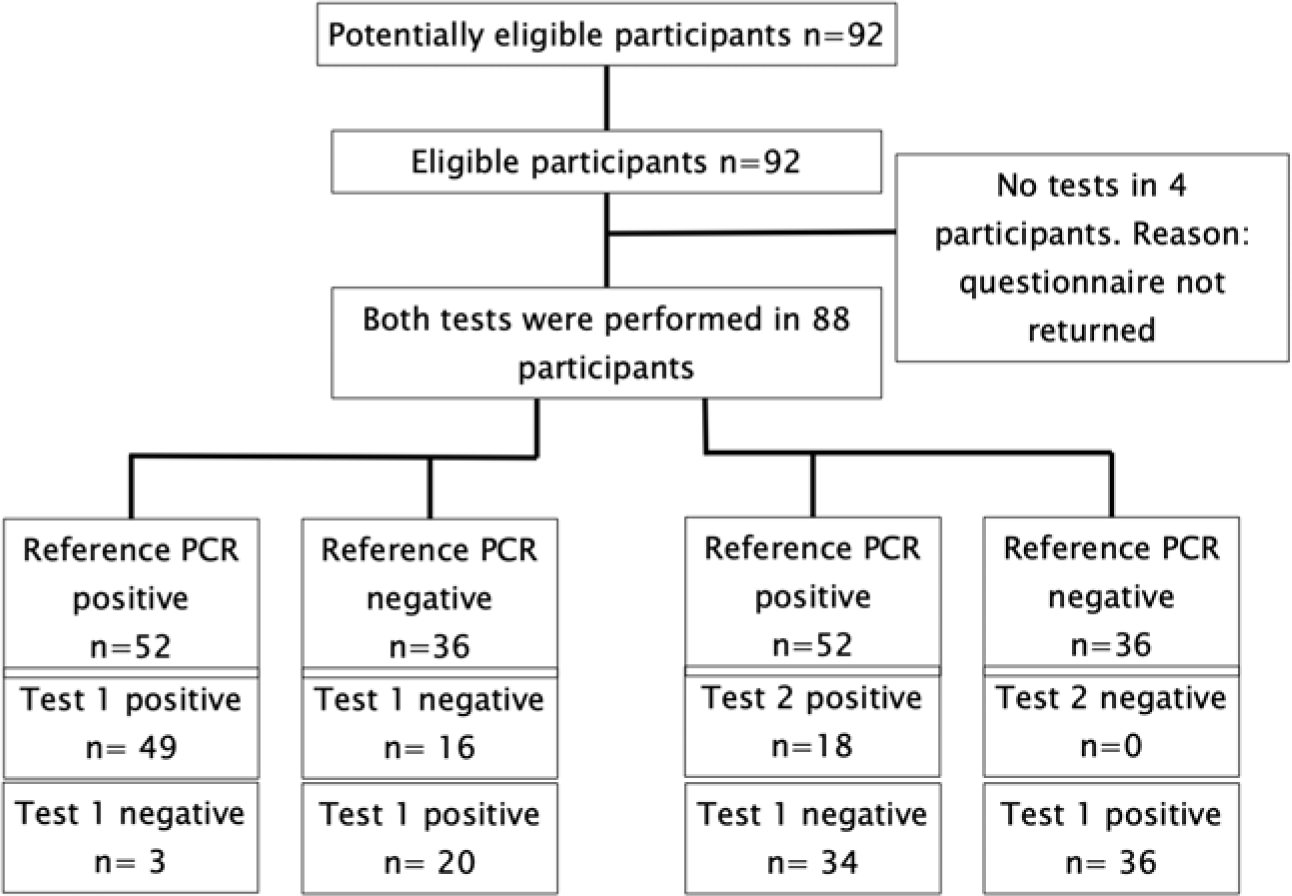
Study flow diagram. Test 1 – sensitivity test: positive questionnaire or/and sweet taste ageusia at a sucrose concentration of 40 mg/l. Test 2 – specificity test: sweet taste ageusia at a sucrose concentration of 106.4 mg/l or/and salty taste ageusia at an NaCl concentration of 13.5 or 17 mg/ml. There were no inconclusive test results.

The study was approved by the proper Ethics Commission for Central Clinical Hospital of the Ministry of the Interior and Administration in Warsaw and was performed in accordance with good clinical practice and the ethical standards laid down in the 1964 Declaration of Helsinki and its later amendments. No external funding or support was available.

A full gustatory function assessment (four flavours) was performed in all subjects. The tastant tablets were prepared in a manner analogous to the pilot study. The tastant tablet was a 0.33 ml gelatine capsule with added flavour. Each flavour capsule was placed in a separate package with a corresponding sample number. Every participant received 10 tablets – one tasteless reference and nine tastant tablets. Neither the patients nor the researchers knew the flavours of the individual samples. The patients were instructed to rinse their mouths with water and to begin testing the flavours with a tasteless sample that was clearly marked and subsequently proceed to the flavour samples (in a random sequence). After tasting each flavour, the participants were instructed to wait until the capsule had dissolved in the mouth and then describe the perceived taste on a form. No adverse events occurred during the test procedures.

### Statistical Analysis

Statistical analyses were conducted with PQStat version 1.8.0.392 and IBM SPSS Statistic version 21. As the entire accessible student population was tested, no prior sample calculation was performed. However, a post-hoc power calculation was performed using a Chi-square test (RxC) for a population of 88 participants. The calculated post-hoc power for each test is shown in Table 3.

There were no undetermined results or missing values. The descriptive statistics for the quantitative variables are given as the mean ± SD. The statistical analysis of different flavour dysfunctions and questionnaire questions between the population subgroups was performed using the chi-square test. If there were fewer than five people in any field of the analysed subgroup, a Yates correction was applied. The level of statistical significance was set at P ≤ .05 with a 95% confidence interval.

The diagnostic test confidence interval (sensitivity, specificity and predictive values of the particular taste) was calculated based on the Clopper–Pearson method for a single proportion. The clinical accuracies of the tests were determined using receiver operator characteristic (ROC) plots. The ROC area under the curve (AUC) was calculated as the fraction ‘correctly identified to be positive’ and the fraction ‘falsely identified to be positive’ determined according to the questionnaire answers and flavour tests.

## Results

The characteristics of SARS-CoV-2 positive patients and SARS-CoV-2 negative students who served as a control group are listed in Table 1. The groups did not differ in terms of their age, height and weight. Almost all participants were male. There were no significant differences concerning the percentage of smokers or vapers in both groups. From the signs and symptoms characteristic of COVID-19 (listed in Table 1), only fever was significantly more frequent in the SARS-CoV-2 positive group, as well as self-reported smell and taste disorders.

**Table 1.** Basal characteristics of study participants. * – significant in the Chi square test at p < 0.005 (with Yates correction p < 0.01), ** – significant in the Chi square test at p < 0.0005 (with Yates correction p < 0.005).

| Parameter | SARS-CoV-2 positive patients (n = 52). Mean (min; max) value. | Control Group (n = 36). Mean (min; max) value. |
| --- | --- | --- |
| Age [years] | 21.7 (19; 26) | 20.8 (19; 24) |
| Male sex [n] | 51 | 34 |
| Height [cm] | 181 (170; 198) | 181 (170; 197) |
| Weight [kg] | 78.5 (60; 112) | 78.5 (62; 93) |
| Time from first positive PCR [days] | 5 (1; 8) | NA |
| Signs and symptoms [last 30 days]: |  |  |
| Fever [n] | 21* | 2 |
| Cough [n] | 15 | 5 |
| Shortness of breath [n] | 8 | 1 |
| Rhinitis and/or sore throat | 22 | 13 |
| Time from first signs/symptoms [days] | no reliable data available | NA |
| Traditional smokers (current) | 4 | 7 |
| Vapers (current) | 3 | 1 |
| Self-reported smell disorders | 25* | 3 |
| Self-reported taste disorders | 32** | 2 |

Subjective smell or taste disturbances were reported by 65% of patients in the test group compared with 8% in the control group. After excluding the least specific testers (Sweet 1 and Sour 1; see Table 2) from the statistical analysis, taste disturbances were diagnosed in 50% of subjects in the test group and 22% in the control group.

**Table 2.** Gustatory disorders in SARS-CoV2 positive and negative patients.

| Parameter | Indication | SARS-Cov2 positive subjects (n) | SARS-CoV2 negative subjects (n) | P |
| --- | --- | --- | --- | --- |
| Sweet 1 (40 mg/ml) | Correct | 15 | 22 | 0.002 |
|  | Incorrect | 37 | 14 |  |
| Sweet 2 (80 mg/ml) | Correct | 43 | 32 | 0.42 |
|  | Incorrect | 9 | 4 |  |
| Sweet 3 (106.4 mg/ml) | Correct | 46 | 36 | 0.034 (with Yates correction 0.092) |
|  | Incorrect | 6 | 0 |  |
| Salty 1 (13.5 mg/ml) | Correct | 46 | 36 | 0.04 (with Yates correction 0.105) |
|  | Incorrect | 6 | 0 |  |
| Salty 2 (17 mg/ml) | Correct | 43 | 36 | 0.034 (with Yates correction 0.092) |
|  | Incorrect | 9 | 0 |  |
| Salty 3 (27 mg/ml) | Correct | 50 | 35 | 0.785 (with Yates correction 0.744) |
|  | Incorrect | 2 | 1 |  |
| Sour 1 (6.25 mg/ml) | Correct | 26 | 24 | 0.12 |
|  | Incorrect | 26 | 12 |  |
| Sour 2 (12.5 mg/ml) | Correct | 44 | 34 | 0.153 (with Yates correction 0.277) |
|  | Incorrect | 8 | 2 |  |
| Bitter 1 (40 mg/ml) | Correct | 42 | 33 | 0.156 |
|  | Incorrect | 10 | 3 |  |

Although a numerical difference was demonstrated between COVID-19 patients and the control group for all flavours, for a single flavour concentration, the difference was only significant with the Sweet 1 tastant tablet (p < 0.002). Detailed data are shown in Table 2.

The diagnostic values of the short medical questionnaire, including the subjective, self-reported loss of taste, self-reported loss of smell and fever in the last month, were calculated. These variables were chosen based on the significant difference of their frequency between the COVID-19 patients and the control group (see Table 1). The questionnaire was regarded as positive when at least one sign or symptom was present within the last 30 days. The sensitivity and specificity of the questionnaire in diagnosing COVID-19 are shown in Table 3. As can be seen, both the specificity and sensitivity were relatively high, but were not sufficiently high to create a reliable screening test (where the sensitivity should exceed 90%) or a diagnostic tool (with a specificity of over 90%).

**Table 3.**
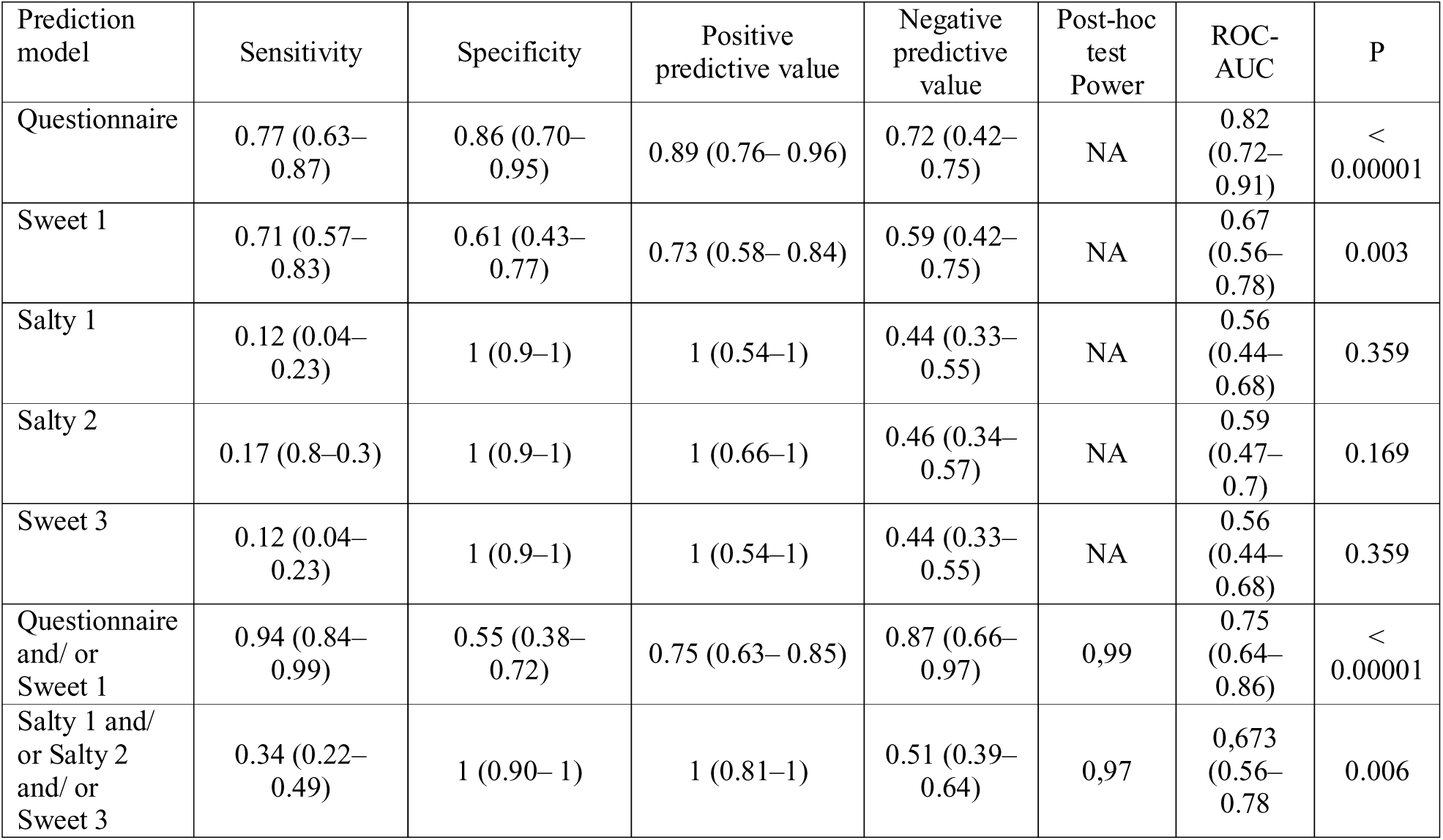
The diagnostic power of the prediction models: specificities, sensitivity, PPV, NPV, ROC-AUC is presented for the sample size n = 88. The questionnaire was positive if at least one of the following signs or symptoms was present: subjective, self-reported loss of taste, self-reported loss of smell, or fever in the last month. NA – not assessed.

| Prediction model | Sensitivity | Specificity | Positive predictive value | Negative predictive value | Post-hoc test Power | ROC-AUC | P |
| --- | --- | --- | --- | --- | --- | --- | --- |
| Questionnaire | 0.77 (0.63–0.87) | 0.86 (0.70–0.95) | 0.89 (0.76–0.96) | 0.72 (0.42–0.75) | NA | 0.82 (0.72–0.91) | < 0.00001 |
| Sweet 1 | 0.71 (0.57–0.83) | 0.61 (0.43–0.77) | 0.73 (0.58–0.84) | 0.59 (0.42–0.75) | NA | 0.67 (0.56–0.78) | 0.003 |
| Salty 1 | 0.12 (0.04–0.23) | 1 (0.9–1) | 1 (0.54–1) | 0.44 (0.33–0.55) | NA | 0.56 (0.44–0.68) | 0.359 |
| Salty 2 | 0.17 (0.8–0.3) | 1 (0.9–1) | 1 (0.66–1) | 0.46 (0.34–0.57) | NA | 0.59 (0.47–0.7) | 0.169 |
| Sweet 3 | 0.12 (0.04–0.23) | 1 (0.9–1) | 1 (0.54–1) | 0.44 (0.33–0.55) | NA | 0.56 (0.44–0.68) | 0.359 |
| Questionnaire and/ or Sweet 1 | 0.94 (0.84–0.99) | 0.55 (0.38–0.72) | 0.75 (0.63–0.85) | 0.87 (0.66–0.97) | 0.99 | 0.75 (0.64–0.86) | < 0.00001 |
| Salty 1 and/ or Salty 2 and/ or Sweet 3 | 0.34 (0.22–0.49) | 1 (0.90–1) | 1 (0.81–1) | 0.51 (0.39–0.64) | 0.97 | 0.673 (0.56–0.78) | 0.006 |

For this reason, additional objective test measures were included to construct different prediction models. Adding the most sensitive tastant – Sweet 1 – to the medical questionnaire produced a model with a sensitivity of over 94% (Table 3 and Figure 2).

**Figure 2.**
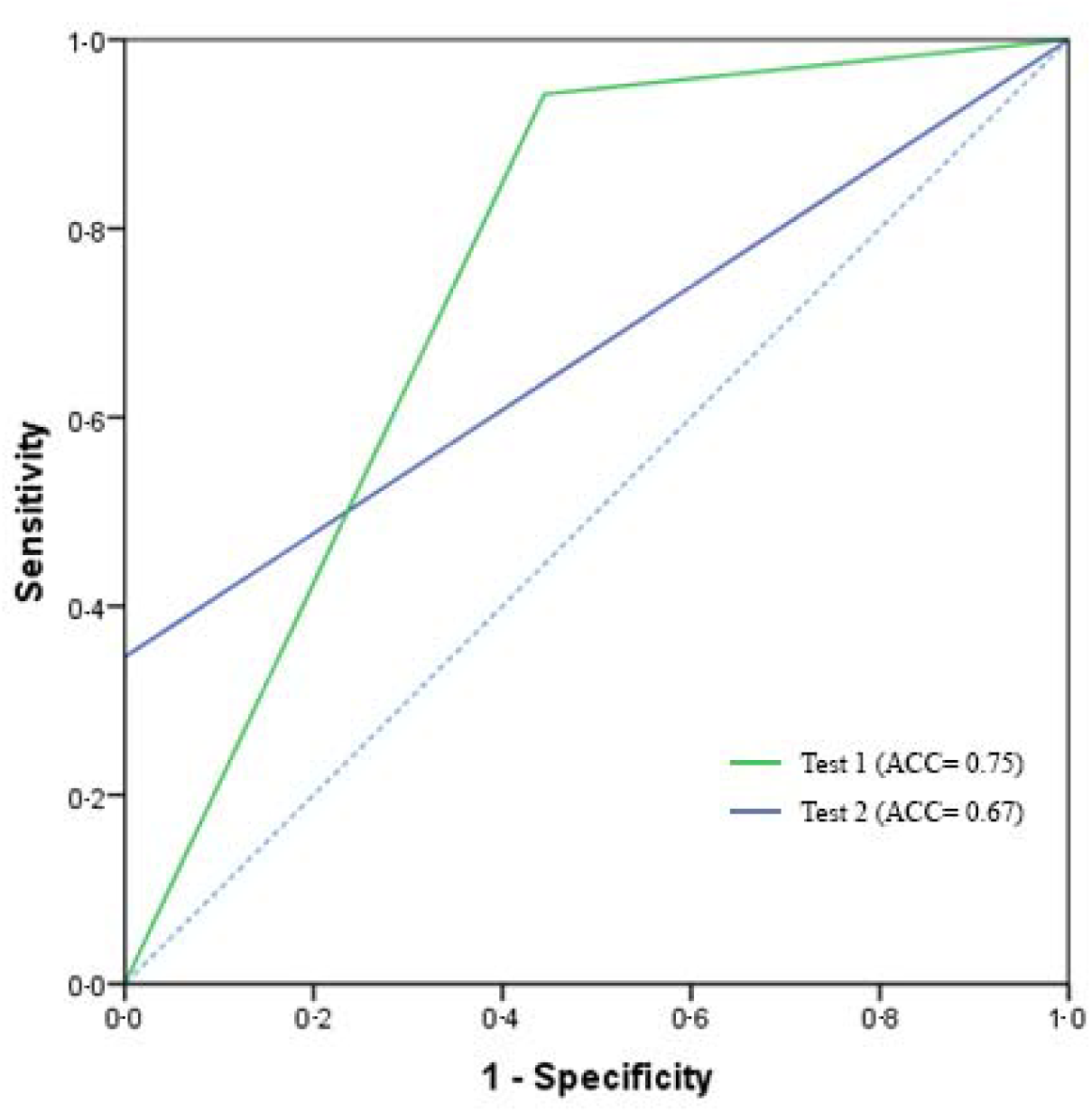
ROC curves for prediction of the risk of a positive test set for SARS-CoV-2. The asymptotic significance of each model was less than 0.05.

However, no model, including the medical questionnaire, was sufficiently sensitive (over 90%). Therefore, a model was created using the most specific tastant tablets (Salty 1, Salty 2, Sweet 3). Although the tablets were not significant separately (Table 3), in parallel testing, the three tastants showed an extremely high specificity of 100%. The diagnostic test evaluation of the different test sets results is shown in Table 3. Figure 2 shows the ROC curves for the prediction models: questionnaire or/and Sweet 1 taste vs Salty 1 or/and Salty 2 or/and Sweet 3.

## Discussion

Recently, taste and smell disturbances have been recognised as symptoms of COVID-19 and have even been listed as key diagnostic factors for COVID-19 by the BMJ.^17,18,19^ Following this discovery, some institutions began to use questionnaires identifying loss of smell and taste as a screening for their visitors.^20^ However, to date, to the best of our knowledge, no study has assessed the accuracy of taste disturbances as a screening tool for COVID-19.

This study confirms and demonstrates that chemosensory taste impairment can be regarded as a symptom of COVID-19. The reason for taste disturbances in COVID-19 is unclear; however, impairment of the renin–angiotensin system may be involved.^21^ In the present study, almost 70% of young, otherwise healthy SARS-COV-2 positive patients reported smell disorder and more than 60% reported taste disorder (Table 1). These figures are about 10 times higher than the matched control group, in which over 8% and almost 6% reported smell disorder and taste disorder, respectively.

This study is, to the best of our knowledge, the first to objectively assess COVID-19-associated taste disturbances and compare their frequency with a control group. It is also the first study to use varying flavour concentrations that were previously validated in healthy subjects. It was shown that the only taste that was significantly more frequently disturbed in SARS-CoV-2 positive patients was the sweet taste at the lowest flavour concentration (40 mg/ml) – a concentration that was perceptible to more than 90% of healthy subjects in a pilot study. This is a unique observation, particularly considering that, until now, reports regarding sweet taste disturbances in COVID-19 patients have been anecdotal.^22^

Other researchers have identified a relatively similar frequency of taste disorders in COVID-19 patients. The highest prevalence of self-reported gustatory disorders identified is 88% in a multicentre European study. This figure is higher than the figure reported in the present research and may be due to the fact that the population described by Lechien et al. also included older subjects (up to 77 years) with concomitant diseases that can also affect taste and cause taste-threshold disturbances.5 No control group was included in the study by Lechien et al. In another study, Yan et al. found self-reported taste disturbances among 71% of SARS-CoV-2 patients and 17% of non-infected American subjects. These results seem to be comparable to ours. Although many of the control subjects in the study by Yan et al. exhibited influenza-like symptoms, including rhinitis and sore throat, which may have increased the frequency of smell and taste disturbances in the group,^6^ the same was true in our study (Tab. 1). Interestingly, the prevalence of dysgeusia reported for European and American populations appears to be much higher than in Chinese SARS-CoV-2 infected patients(5.6%), as reported by Mao et al,^23^ and Korean patients (15.3%) as reported by Lee et al.^24^

One study that objectively assessed gustatory dysfunction in COVID-19 patients only used single concentrations of particular flavors.^11^ The patients in this study were considered to have ageusia if they were not able to identify any of the following four tastes: sweet, salty, sour and bitter. The frequency of ageusia according to this definition was 1.4% of study subjects, while 51.4% accurately identified all tastes. However, no control group was included in this study; in light of this, and due to differences in flavour preparation and concentrations, it is difficult to compare the results with those of the present study (although the figure of 51.4% is similar to the 50% of patients in the present study who accurately identified all tastes after the exclusion of Sweet 1 and Sour 1).

We do not consider the objective taste measurement (even the disordered sweet taste at a concentration of 40 mg/ml, which had a sensitivity of 71%; see Table 3) sufficient as a screening test for COVID-19. However, when this test was combined with a questionnaire consisting of three questions (whether patients had smell or taste disorders or fever in the last 30 days), the sensitivity increased to 94%. This appears to be an entirely acceptable value, taking into account that, for example, the sensitivity of the commonly used PSA screening of prostate cancer may be as low as 33% for a cut-off level of 3 ng/ml, sensitivity that makes this screening not reliable.^25,26^ Our predictors appear to be similar to those found by Menni et al. in a large population sample, although these authors only utilised self-reported data, and the sensitivity and specificity (66% and 78% in a UK population, respectively) of the proposed model was much lower than ours.^27^

A 94% sensitivity test, particularly if simple, inexpensive and safe, may be used for disease screening in large populations. Such a test may be also used repeatedly (e.g., once a week) to ensure fast identification of at-risk subjects and could be performed as a point-of-care test or as a self-administered test. Although we did not compare the results of the self-administered vs operator-administered test, a comparison with another, more complicated test showed no significant differences between the two procedures.^28^ Such a test could also be used in developing countries in which the availability of genetic (RT-PCR for SARS-CoV-2) or serological (antibodies) screening is low. It could be used also in large companies or plants, where the need for nasopharyngeal swabs or blood draw, particularly when performed frequently, as well as the requirement to transport samples, may represent a significant burden.

The specificity of the proposed test is 55% (Table 3), which means that around as many as 50% of identified subjects will have COVID-19. This appears to be acceptable for a screening test, though the value is too low for it to be used as a reliable diagnostic tool. However, a test that combines three tastants (Salty 1 or Salty 2 and/or Sweet 3) demonstrated 100% specificity in diagnosing COVID-19. A positive result of such a test in asymptomatic or oligosymptomatic patients could serve as a basis for referring patients to quarantine in the event of limited availability of genetic or serological tests, as is the case in many countries in the world. It should be remembered that an estimated 50–70% of active cases may be asymptomatic at the time of diagnosis.^15^

This study has some limitations. A relatively low number of patients implies a validation in the bigger population, which is planned in the future. However, one of the strengths of this study is that it was performed on a uniform, young and otherwise healthy population living in a similar environment, with a comparable examined and control group.

The fact that the majority of subjects in the examined and control group in this study were men can be regarded as a limitation. However, as SARS-CoV-2 positive men seem to have gustatory dysfunction more rarely that women, the results could be even more reliable in women and in the general population.^5^ Another limitation is the limited accuracy of RT-PCR tests, which are the gold standard of COVID-19 diagnosis. To confirm the results of a PCR test, each patient underwent SARS-CoV-2 tests twice. In one case, a patient who initially received a negative diagnosis subsequently obtained a confirmed positive result.

In summary, the pandemic situation not only checks the state’s health security system but above all affects the sense of security of the individual. Regardless of psychophysical resilience, it destabilizes functioning, especially in the absence of information regarding potential infection. Hence, tests, especially those widely available, are crucial in satisfying the basic human needs related to their health safety.

At present, the gold standard for the detection of SARS-CoV-2 infection, according to the WHO, is RT-PCR examination of nasopharyngeal swab. However, the sensitivity of RTPCR as a screening method can be reduced in the case of incorrectly performed swabs. The costs of this method as a screening test is extremely high, the logistics involved in providing samples to reference laboratories is complicated, and the waiting time for results can be very long. Thoracic CT scan, on the other hand, demonstrates high sensitivity and specificity for COVID-19. However, in the context of screening, Thoracic CT scan also carries a number of disadvantages, including a high x-ray load during the procedure. Both methods are not accessible to the wider population in numerous countries.

As a SARS-CoV-2 vaccine is not yet available, and with only one vaccine candidate currently in a phase 2 trial, the only method that can currently control the pandemic involves testing the broader population for symptomatic, oligosymptomatic and asymptomatic carriers of SARS-CoV-2, isolating infected subjects and initiating early treatment of patients who develop COVID-19 symptoms.^29^ An inexpensive, simple, fast and sensitive screening method is needed for this purpose. There is currently a public debate on the introduction of this type of test.^30^ Considerable attention was paid to population testing strategies in the latest edition of the Lancet Infectious Diseases.^31^ Before sample-in-answer-out systems are introduced on a large scale, particularly in public utilities, and mass screening is introduced in developing countries, a system based on flavour tests could improve COVID-19 screening by identifying possible carriers of SARS-CoV-2 for further serological, molecular or other testing. Such a test, of the kind proposed in this study, could also be used to diagnose SARS-CoV-2 infection and serve as a basis for referring patients to quarantine in the event of limited availability of genetic or serological tests.

## Data Availability

All available data are accesible in the manuscript.

